# Risk Factors, Clinical Outcomes and Prognostic Factors of Bacterial Keratitis: The Nottingham Infectious Keratitis Study

**DOI:** 10.1101/2021.05.26.21257881

**Authors:** Darren S. J. Ting, Jessica Cairns, Bhavesh P. Gopal, Charlotte Shan Ho, Lazar Krstic, Ahmad Elsahn, Michelle Lister, Dalia G. Said, Harminder S. Dua

**Author notes:** **Corresponding author:** Professor Harminder S. Dua; **Email:**, **Corresponding address:** Academic Ophthalmology, Division of Clinical Neuroscience, School of Medicine, University of Nottingham, Nottingham, NG7 2RD, UK. Equally contributing authors (Dalia G. Said, Harminder S. Dua).

## Abstract

**Background/aim:** To examine the risk factors, clinical characteristics, outcomes and prognostic factors of bacterial keratitis (BK) in Nottingham, UK.

**Methods:** This was a retrospective study of patients who presented to the Queen’s Medical Centre, Nottingham, with suspected BK during 2015-2019. Relevant data, including the demographic factors, risk factors, clinical outcomes, and potential prognostic factors, were analysed.

**Results:** A total of 283 patients (n=283 eyes) were included; mean age was 54.4±21.0 years and 50.9% were male. Of 283 cases, 128 (45.2%) cases were culture-positive. Relevant risk factors were identified in 96.5% patients, with ocular surface diseases (47.3%), contact lens wear (35.3%) and systemic immunosuppression (18.4%) being the most common factors. Contact lens wear was most commonly associated with *P. aeruginosa* whereas *Staphylococci spp*. were most commonly implicated in non-contact lens-related BK cases (p=0.017). At presentation, culture-positive cases were associated with older age, worse presenting corrected-distance-visual-acuity (CDVA), larger epithelial defect and infiltrate, central location and hypopyon (all p<0.01), when compared to culture-negative cases. Hospitalisation was required in 57.2% patients, with a mean length of stay of 8.0±8.3 days. Surgical intervention was required in 16.3% patients. Significant complications such as threatened/actual corneal perforation (8.8%), loss of perception of light vision (3.9%), and evisceration/enucleation (1.4%) were noted. Poor visual outcome (final corrected-distance-visual-acuity of <0.6 logMAR) and delayed corneal healing (>30 days from initial presentation) were significantly affected by age >50 years, infiltrate size >3mm, and reduced presenting vision (all p<0.05).

**Conclusion:** BK represents a significant ocular morbidity in the UK. Culture positivity is associated with more severe disease at presentation but has no significant influence on the final outcome. Older age, large infiltrate, and poor presenting vision were predictive of poor visual outcome and delayed corneal healing, highlighting the importance of primary prevention and early intervention for BK.

## INTRODUCTION

Infectious keratitis is a major cause of corneal blindness in both developed and developing countries.^1^ The incidence has been estimated at 2.5-799 cases per 100,000 population/year.^1-4^ Subject to geographical, temporal and seasonal variations, bacteria and fungi are the most commonly implicated organisms in infectious keratitis.^4-8^ The variations are mainly attributed to the difference in the climate of the studied region and the population-based risk factors, particularly contact lens wear, trauma, and agricultural activities.

Bacterial keratitis (BK) has been consistently shown to be the main causative organisms in the UK and other developed countries. Based on the recent literature, BK represents 90-93% and 72-86% of all culture-proven infectious keratitis cases in the UK and in North America, respectively.^4,9-13^ In our recent Nottingham Infectious Keratitis Study, we observed that 92.8% of the culture-proven infectious keratitis cases were caused by bacteria, with *Pseudomonas aeruginosa* being the most common isolate.^4^ In addition, we observed a seasonal predilection of *P. aeruginosa* keratitis in summer, which has been hypothetically linked to the increased use of contact lens wear and trauma during outdoor/water activity,^6^ though such association remains to be elucidated.

In view of the prevalence of BK in the UK and other parts of the world, it is important to understand the underlying risk factors (for preventative measures) and the clinical outcomes of BK. To date, the majority of UK studies had largely focussed on the epidemiology, causative microorganisms and antimicrobial resistance of bacteria,^4,9-11^ with limited information on the risk factors and outcomes of BK.^14,15^ In this study, we aimed to examine the risk factors, clinical characteristics, outcomes and prognostic factors of BK in Nottingham, UK.

## MATERIALS AND METHODS

This was a retrospective study of all cases of BK that presented to the Queen’s Medical Centre, Nottingham, UK, between January 2015 and December 2019 (a 5-year period). The study was approved by the Clinical Governance team in the Nottingham University Hospitals NHS Trust as a clinical audit (Ref: 19-265C). Ethical approval was not required as this study involved retrospective examination of the medical case notes and did not involve any human participants. Similarly, written informed consent from the patients was not required to participate in this study in accordance with the national legislation and the institutional requirements.

### Case identification and definition

Potential cases of BK were first identified via the local microbiological database as described in previous studies.^4,6^ Subsequently, the medical case records were examined to confirm the eligibility of the potential cases prior to inclusion into the study. Both culture-positive and culture-negative presumed BK cases were included in this study. Culture-positive BK was defined as the presence of clinical BK with confirmation of the causative bacteria on microbiological culture. Culture-negative BK was diagnosed based on the clinical findings and the clinical course of the disease where improvement and/or resolution of the infection was achieved by intensive topical antibiotic treatment without other types of antimicrobial treatment. Cases that did not have complete initial and/or follow-up data were excluded from this study. Fungal, viral and parasitic keratitis were excluded from this study.

### Data collection

Relevant data, including demographic factors, risk factors, clinical characteristics, types of bacteria, corrected-distance-visual-acuity (CDVA), management, outcome and complications, were collected using a standardised excel proforma. Risk factors were divided into: (1) contact lens wear; (2) trauma; (3) ocular surface diseases (e.g. dry eye, meibomian gland dysfunction, neurotrophic keratopathy, exposure keratopathy, previous corneal infection, recurrent corneal erosion syndrome, limbal stem cell deficiency, cicatricial conjunctivitis, band keratopathy, and bullous keratopathy); (4) use of topical corticosteroids; (5) previous/recent history of corneal surgery (e.g. corneal graft, pterygium surgery, corneal collagen cross-linking and corneal debridement/delamination); and (6) systemic immunosuppression (e.g. diabetes, systemic immunosuppressive treatment, malnutrition, and immunodeficiency). The size of epithelial defect and infiltrate were categorised as small (<3mm), moderate (3.1-6mm), or large (>6mm), based on the maximum linear dimension (**Figure 1A-C**). The location of the ulcer was divided into peripheral (the entire ulcer was within 3mm from the limbus, paracentral (in between the central and peripheral location), and central (any part of the ulcer affecting the visual axis; **Figure 1D-F**). Recurrence was defined as the re-occurrence of BK after complete resolution of the previous BK episode, irrespective of the time interval between the first and subsequent infective episode. To avoid any duplication of the patient’s risk factors in bilateral or recurrent BK cases, we only included one eye per patient in this study. For recurrent cases, only the first BK episode was included and analysed, regardless of the laterality of infection in the subsequent infective episode.

**Figure 1.**
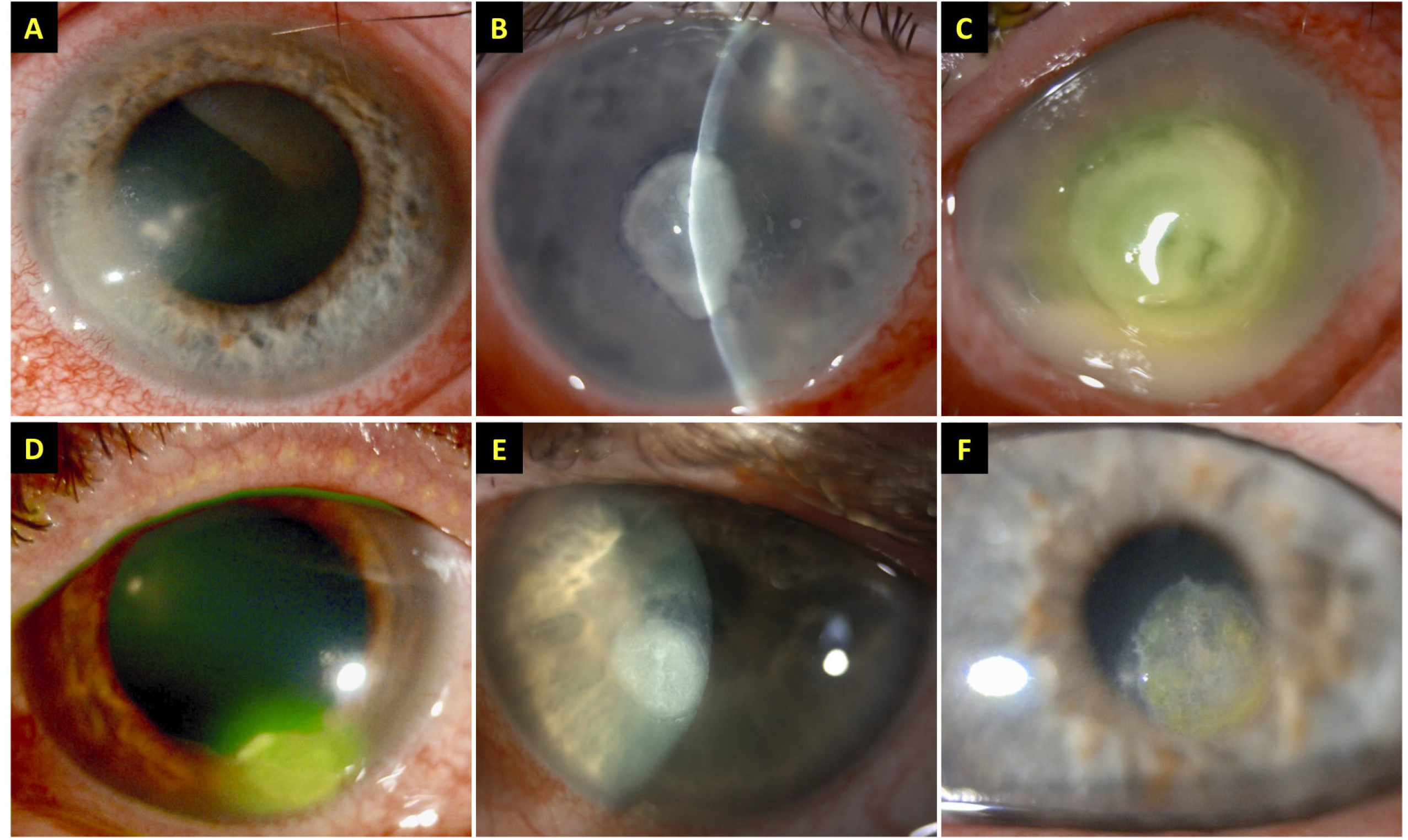
**(A-C)** Examples of bacterial keratitis with varying severity, including **(A)** small infiltrate (<3mm), **(B)** moderate infiltrate (3.1-6mm), and **(C)** large infiltrate (>6mm). **(D-F)** Examples of bacterial keratitis in different locations, including **(D)** peripheral, **(E)** paracentral, and **(F)** central location.

### Microbiological culture, diagnosis and treatment

Based on the departmental guideline for infectious keratitis, all patients presented with corneal ulcer(s) of >1 mm diameter, central location, associated with significant anterior chamber reaction/hypopyon, or atypical presentation were subjected to microbiological investigation with corneal scraping for microscopy (with Gram staining) and microbiological culture and sensitivity testing.^4,6^ Corneal scrapes were inoculated on chocolate agar (for fastidious organisms), blood agar (for bacteria), and Sabouraud dextrose agar (for fungi). For suspected cases of Acanthamoeba keratitis, corneal swab and/or epithelial biopsy was obtained for culture on non-nutrient agar with *Escherichia coli* overlay. All cultures were incubated for at least 1 week (and up to 3 weeks for suspected Acanthamoeba keratitis and fungal keratitis). The identity of microorganisms was confirmed through standard culture and bacteriology tests. In vivo confocal microscopy (IVCM) using the Heidelberg Retinal Tomography (HRT) II with Rostock Cornea Module (Heidelberg Engineering Ltd, Hertfordshire, UK) was utilised to aid the diagnosis (or exclusion) of fungal and Acanthamoeba keratitis.

All patients with BK were started on hourly topical treatment using either levofloxacin 0.5% monotherapy or combined therapy of fortified cephalosporin (cefuroxime 5%) and aminoglycoside (either amikacin 2.5% or gentamicin 1.5%), based on the severity of cases and the clinician’s preference. Hospitalisation was warranted if the ulcer was severe (i.e. central, infiltrate >2mm, or presence of hypopyon) or was unresponsive to the initial antibiotic treatment, or the patient was unable or unlikely to comply with the intensive treatment regimen. All patients that were admitted for treatment were started on the combined therapy. Further changes to the antibiotic treatment were made, if necessary, based on the clinical course and the microbiological results.

### Statistical analysis

Statistical analysis was performed using SPSS version 26.0 (IBM SPSS Statistics for Windows, Armonk, NY, USA). For descriptive and analytic purposes, the cases were divided into culture-positive and culture-negative BK cases. Comparison between groups was conducted using Pearson’s Chi square or Fisher’s Exact test where appropriate for categorical variables, and T test or Mann-Whitney U test for continuous variables. All continuous data were presented as mean ± standard deviation (SD) and/or 95% confidence interval (CI).

The main outcome measures were corrected-distance-visual-acuity (CDVA) and time to complete corneal healing, defined as complete resolution of infection with corneal re-epithelialisation. Snellen vision was converted to logMAR vision for analytic purpose. Vision of counting fingers (CF), hand movement (HM), perception of light (PL) and no perception of light (NPL) were quantified as 1.9 logMAR, 2.3 logMAR, 2.8 logMAR and 3.0 logMAR respectively.^16,17^ For cases that required therapeutic or tectonic keratoplasty, the vision prior to the transplant was used as the final CDVA. In enucleation or evisceration cases, a CDVA of 3.0 logMAR (equivalent to NPL vision) was assigned as the final vision. Multivariable logistic regression analysis was performed to examine for any potential prognostic factors for poor visual outcome, defined as CDVA of worse than 6/24 (or <0.6 logMAR), and poor corneal healing, defined as >30 days to achieve complete corneal healing, occurrence of uncontrolled infection or corneal perforation requiring corneal gluing, tectonic or therapeutic keratoplasty, and/or evisceration / enucleation. The results of logistic regression analyses were presented in odd ratios (ORs) with 95% confidence interval (CI). P-value of <0.05 was considered statistically significant.

## RESULTS

### Overall description

During the 5-year study period, a total of 283 patients (n = 283 eyes) with BK were included. The mean age was 54.4 ± 21.0 years (range, 4.9-92.7 years), 50.9% patients were male, and 51.2% cases affected the left eye (**Table 1**). Two bilateral BK cases were identified and only the right eye was included. The mean follow-up duration was 6.0 ± 8.9 months. Of all included cases, 128 (45.2%) and 155 (54.8%) cases were culture-positive and culture-negative BK (**Table 1)**.

### Risk factors and causative organisms

Nearly all (273, 96.5%) patients were found to have at least one risk factor, with 66 (23.3%) patients having two risk factors, and 18 (6.4%) patients having three or more risk factors for BK. Ocular surface diseases (134, 47.3%) were the most common risk factor, followed by contact lens wear (100, 35.3%), systemic immunosuppression (52, 18.4%), prior corneal surgery (39, 13.8%), use of topical corticosteroids at presentation (31, 11.0%), and trauma (25, 8.8%; **Table 1**). Contact lens wear was more commonly associated with younger patients (≤50 years) whereas systemic immunosuppression was more commonly associated with older patients (>50 years; **Table 2**).

Of the 128 culture-positive cases, 10 (7.8%) cases grew more than one species, with a total of 138 bacteria being identified (**Table 3**). *Pseudomonas aeruginosa* (44, 31.9%) was the most common isolate identified, followed by *Staphylococci spp*. (36, 26.1%) and *Streptococci spp*. (16, 11.6%). Contact lens wear was most commonly associated with *P. aeruginosa* (23, 51.1%) whereas *Staphylococci spp*. were most commonly implicated in non-contact lens-related BK cases, including those affected by ocular surface disease (20, 31.7%), previous history of corneal surgery (10, 40.0%), and use of topical corticosteroids (7, 33.3%; p=0.017; **Table 3**).

### Clinical characteristics

The baseline clinical characteristics are summarised in **Table 1**. At baseline, 124 (43.8%) patients presented with a CDVA of <1.0 logMAR. The most commonly observed clinical characteristics of the ulcer were small epithelial defect size (172, 60.8%), small infiltrate size (183, 64.7%), central location (110, 38.9%), and absence of hypopyon (201, 71.0%). The mean duration of symptoms prior to presentation was 6.0 ± 13.0 days. Hospitalisation for intensive treatment was required in 162 (57.2%) patients, with a mean hospitalisation duration of 8.0 ± 8.3 days. The baseline clinical characteristics of BK were significantly different between culture-proven and culture-negative cases. Culture-proven cases were more commonly associated with older age (p=0.004), prior corneal surgery (p=0.011), use of topical corticosteroids (p=0.008), poorer presenting CDVA (p<0.001), larger epithelial defect / infiltrate size (p<0.001), central or paracentral ulcer (p=0.002), presence of hypopyon (p<0.001), and need for hospitalisation for intensive treatment (p<0.001).

### Medical and surgical treatment

A total of 237 (83.7%) patients were successfully treated with medical treatment alone, while 46 (16.3%) patients required additional surgical interventions for controlling the infection and/or its sequelae. Various surgical interventions were performed, including corneal gluing (22, 7.8%), temporary / permanent tarsorrhaphy (13, 4.6%), single or multi-layer amniotic membrane transplant (11, 3.9%), conjunctival hooding (3, 1.1%), and emergency therapeutic / tectonic keratoplasty (2, 0.7%), evisceration (2, 0.7%), and enucleation (2, 0.7%). Five (1.8%) patients required elective optical penetrating keratoplasty after the resolution of infection.

### Clinical outcomes and prognostic factors

The mean CDVA (in logMAR) improved from 1.17 ± 1.03 at presentation to 0.80 ± 1.00 at final follow-up (p<0.001). From baseline to final follow-up, the proportion of patients with CDVA of ≥0.30 logMAR improved from 31.4% to 53.0%, with the proportion of CDVA of <1.0 logMAR reducing from 43.8% to 30.4% (p<0.001; **Figure 2**). Twenty-three (8.1%) patients had a final CDVA of PL or worse, including four (1.4%) patients that had evisceration / enucleation. Multivariable logistic regression demonstrated that poor visual outcome (CDVA <0.6 logMAR) was significantly influenced by age >50 years old (OR 2.61; 95% CI, 1.24-5.47; p=0.011), infiltrate size >3mm (OR 4.07; 95% CI, 1.21-13.73; p=0.024), central ulcer (OR 2.13; 95% CI, 1.01-4.51; p=0.047), and presenting CDVA of <0.6 logMAR (OR 29.70; 95% CI, 10.47-84.18; p<0.001; **Table 4**).

**Figure 2.**
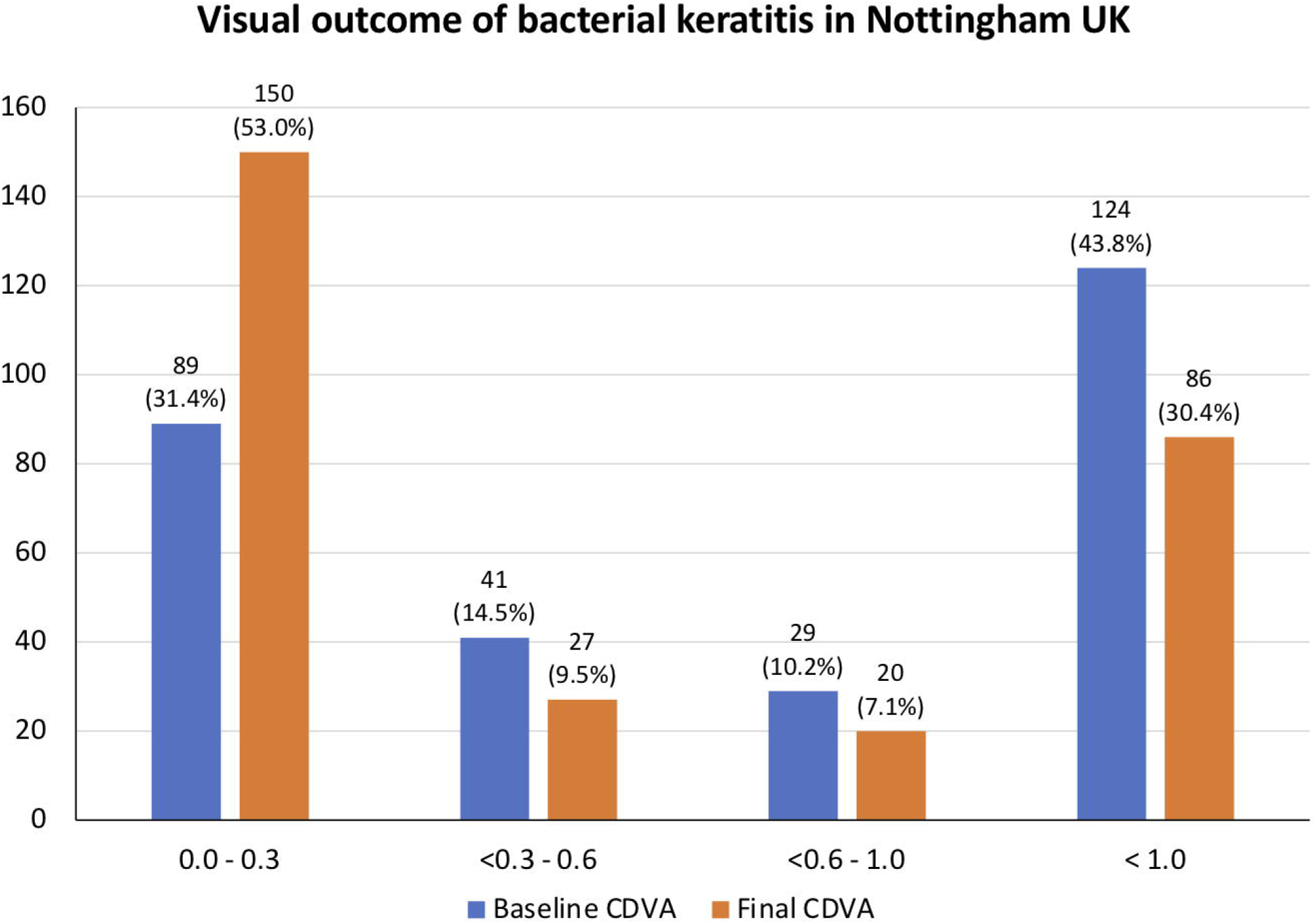
Summary of corrected-distance-visual-acuity (CDVA) of patients with bacterial keratitis at initial presentation and at final follow-up.

In terms of complete corneal healing, 278 (98.2%) patients achieved complete corneal healing at final follow-up, with four patients requiring evisceration / enucleation and one patient was still undergoing active treatment. The mean corneal healing time was 1.6 ± 1.5 months, with 157 (55.5%) patients having a corneal healing time of >30 days. Multivariable logistic regression analysis demonstrated that poor corneal healing (>30 days to achieve complete healing) was significantly affected by age >50 years (OR 1.86; 95% CI, 1.06-3.24; p=0.030), involvement of right eye (OR 1.82; 95% CI, 1.05-3.16; p=0.033), infiltrate size >3mm (OR 3.46; 95% CI, 1.24-9.70; p=0.018), and presenting CDVA of <0.6 logMAR (OR 2.22; 95% CI, 1.19-4.15; p=0.013; **Table 4**). Other factors such as gender, culture positivity, and presence of hypopyon did not significantly influence the visual outcome or the corneal healing time (all p>0.05).

### Complications

A number of complications were observed in this study, including raised intraocular pressure (>21 mmHg) / glaucoma (32, 11.3%), recurrence of infection (28, 9.9%), threatened / actual corneal perforation (25, 8.8%), complete loss of vision / NLP (11, 3.9%), loss of eye (4, 1.4%), and phthisis bulbi (1, 0.4%).

## DISCUSSION

BK is the most common cause of infectious keratitis in the UK and in many developed countries. To the best of our knowledge, this study represents one of the largest and most up-to-date studies in the UK specifically examining the risk factors, clinical characteristics, outcomes and prognostic factors of BK.

### Risk factors and causative organisms

BK rarely occurs in the absence of any predisposing factor. In this study, relevant risk factors were identified in 96.5% of the patients, with 29.7% of them having two or more risk factors. Identification of risk factors for BK is important as it allows the clinicians to manage the identified risk factors to reduce the risk of recurrence of infection and helps provide some insights into the underlying causative organisms, thereby guiding the choice of antimicrobial treatment, especially in the absence of positive microbiological culture results.

Risk factors for infectious keratitis have been shown to vary considerably across different studies.^1^ Contact lens wear is one of the most common risk factors for infectious keratitis in the developed countries whereas trauma is most commonly implicated in developing countries.^1,18-20^ Kaye et al.^14^ previously conducted a multi-center study in the UK examining the risk factors and outcomes of BK in 2003-2006. The most common risk factor was found to be corneal / ocular surface diseases (50%) and contact lens wear (32%). Another UK study conducted by our group in 2007-2010 examining the profile of sight-threatening infectious keratitis in Nottingham (which included BK and Acanthamoeba keratitis) observed that ocular surface disease (33%), contact lens wear (26.5%) and previous ocular surgery (20.2%) were the most common risk factors. A similar distribution of the risk factors was observed in our study where ocular surface diseases and contact lens wear were found to be most common factors, suggesting that the population-based risk factors for BK in the UK had remained similar over the past two decades.

In contrast, Khoo et al.^19^ had recently examined the clinical characteristics of infectious keratitis (all types of organisms) in Sydney, Australia, and reported that contact lens wear (63%) and topical use of steroid (24%) were the most common risk factors, highlighting the difference in risk factors among different regions, even in the setting of developed countries. Gaining a better knowledge of the region-specific population risk factors enables a more targeted preventative strategy and research focus for reducing the incidence and burden of infectious keratitis. Interestingly, an immunosuppressed state (which included diabetes) was found to be the third most common risk factor for BK in our study. This might be attributed to the overall reduced immunity and specifically at the ocular surface, promotion of microbial growth (in hyperglycaemia), and presence of undiagnosed ocular surface diseases such as dry eye disease and neurotrophic keratopathy that are commonly linked to diabetes.^1,21,22^

We observed that contact lens-related BK was most commonly caused by *P. aeruginosa*, which is consistent with the findings of many other studies. This observation also provides support to our previous hypothesis on the increased prevalence of *P. aeruginosa*-related BK during the summer season due to increased contact lens wear.^6^ On the other hand, Gram-positive bacteria, including *Staphylococci spp*., which are common ocular surface commensals, are more frequently identified in BK cases affected by ocular surface diseases, prior ocular surgery and use of topical steroids. This was consistent with other studies whereby Gram-positive bacteria were most commonly implicated in BK associated with ocular surface diseases.^23^ Therefore, in non-contact lens-related culture-negative BK cases that are not responsive to fluoroquinolone monotherapy, adding a cephalosporine would be beneficial as it normally provides good coverage to Gram-positive bacteria.^4^

### Clinical characteristics

In our study, we observed that many of the BK cases were of mild severity (i.e. small ulcer size without the presence of hypopyon). This was likely attributed to the fact that the majority of our patient sought medical attention within the first week of their ocular symptoms. This may also explain the lesser (16%) need for additional surgical interventions. On the other hand, an Indian study of infectious keratitis conducted two decades ago showed that only 0.02% of their patients presented within the first week of ocular symptoms, with 12% of the cohort presenting one month after the onset of symptoms. Notably, 43% of their BK patients required surgical interventions, considerably higher than our study. Another recent Indian study of infectious keratitis conducted at another region observed that the median duration of ocular symptoms was 7 days, with 72% cases caused by corneal trauma. The heterogeneity in the promptness of patients seeking medical attention is likely related to the difference in the culture, level of education and health awareness, causes / risk factors (earlier in trauma-related cases), and accessibility to healthcare facility. Although our analysis showed that patients with duration of ocular symptoms of ≥7 days had a worse visual outcome (<0.6 logMAR CDVA), the association was not significant in the multivariable regression analysis (not presented herein). We had not included the duration of ocular symptoms as one of the independent variables in our current regression model in view of the high amount (∼15%) of missing data in this parameter, which could negatively affect the multivariable regression analysis.

### Microbiological culture results and associations

Corneal scraping for culture and sensitivity testing remains the most common microbiological investigations for infectious keratitis. While the culture yield has been shown to be variably low (23.7-77%),^1,24,25^ this is currently the only method that could provide both the information of the underlying causative organisms and the antimicrobial susceptibility and resistance results. In this study, 45% of our cases were culture-proven but this was not truly reflective of the culture yield of infectious keratitis in our region as cases with incomplete data or inconclusive cause were excluded from this study. We observed that culture positivity was significantly associated with several factors, including increased age, large ulcer size, central ulcer, prior corneal surgery or use of topical steroids, and worse presenting vision. Such association is likely attributed to the more severe disease and higher microbial load at presentation. This is in accordance with the “1, 2, 3 Rule” advocated by Vital et al. that corneal culture should be performed when any of the three clinical parameters is met (i.e. ≥1+ anterior chamber cells, ≥2 mm infiltrate, or infiltrate ≤3 mm distance from the corneal center) as it predicts the severity, outcome and likelihood of positive culture in infectious keratitis.^26,27^ In addition, Cariello et al.^28^ similarly showed that previous use of topical steroids increased the chance of positive culture. Therefore, these findings suggest that performing corneal culture in older patients with more severe disease and with prior use of topical steroids is more likely to have a positive culture.

### Outcomes and prognostic factors

The majority (84%) of our cases healed with medical treatment alone. While 25 (9%) patients developed threatened / actual perforation, most of them (21, 84%) were amenable to corneal gluing, multi-layer amniotic membrane transplant or conjunctival hooding, with only 2 (0.7%) requiring emergency tectonic keratopathy. This is in contrast with the findings of the Asia Cornea Society Infectious Keratitis Study (ASCIKS) whereby ∼10% of the cohort required emergency therapeutic keratoplasty.^5^ Another Australian study, which included all types of infectious keratitis, showed that 6% of the patients required either therapeutic keratoplasty, evisceration or enucleation.^19^ The discrepancy among the studies may be related to the difference in the severity of the presenting ulcer, the risk factors (lower proportion of trauma in our study), and the inclusion of fungal keratitis and/or polymicrobial keratitis, which are often more difficult to manage compared to BK.^5,18,19,29,30^

A number of important prognostic factors for visual outcome and corneal healing were identified in our study. We observed that poor visual outcome was significantly influenced by older age, larger infiltrate, central ulcer, and poor presenting CDVA. Khoo et al.^19^ similarly demonstrated that older patients with worse presenting vision and larger ulcer were more likely to experience a poor outcome, which was defined as vision of <6/60, decrease vision during treatment, or occurrence of complications requiring keratoplasty, evisceration or enucleation. Parmar et al.^31^ also reported that elderly patients (≥65 years old) were more commonly affected by central and larger ulcers and worse visual outcome.

Additionally, we showed that corneal healing was negatively affected by older age, larger infiltrate size and poorer presenting vision. Gaining a better knowledge of these prognostic factors may enable earlier interventions (e.g. earlier use of regular lubricants, temporary tarsorrhaphy or amniotic membrane transplant) to help promote corneal healing and re-epithelialisation after the acute sterilisation phase.^32-34^ The poorer corneal wound healing in older patients with BK is likely related to the presence of co-existing ocular surface diseases (e.g. dry eyes, neurotrophic keratopathy, and others), immunosuppression and the age-related reduction in proliferative ability of the limbal stem cells.^35-37^

### Strengths and limitations

This study serves as one of the largest and most up-to-date examination of the risk factors, clinical characteristics and outcomes of BK in the UK. One of the limitations of this study was the inclusion of culture-negative BK cases. However, we had examined the medical case notes to ensure that these cases were true BK cases based on the clinical presentation and the clinical course. In addition, inclusion of the culture-negative cases enabled the examination of potential predictive factors for culture positivity and the outcome of these cases as culture-negative BK cases represents a large proportion of infectious keratitis in clinical practice. The issue with low culture yield in infectious keratitis has been consistently highlighted in many studies.^1,2^ To overcome this clinical barrier, a number of novel and emerging technologies, including MALDI-TOF mass spectrometry,^38,39^ IVCM,^40^ polymerase chain reaction (PCR),^41^ next generation sequencing,^42^ and artificial intelligence-assisted platforms,^43^ have been developed and/or implemented in clinical practice. In addition, emerging treatment such as therapeutic corneal collagen cross-linking, ultraviolet C treatment, polymer-based treatment and antimicrobial peptides may serve as useful adjunctive treatment in the near future, improving the management and treatment outcome of infectious keratitis.^44-48^

In conclusion, BK represents a significant ocular morbidity in the UK. It not only significantly affects the patients’ vision but also places considerable burden on the healthcare services as hospital admission is often required for intensive medical treatment and/or surgical intervention for BK. Affected patients are usually working adults (18-64 years) and hence the disease can have significant impact on the public and private workforce. As the visual outcome of BK is affected by the initial severity of the infection and the presenting vision, the importance of “prevention is better than cure” cannot be overemphasised. Ocular surface diseases, contact lens wear and systemic immunosuppression are important risk factors for BK and better preventative strategies need to be developed and targeted towards these areas. Future studies evaluating the risk factors and outcomes of other types of infectious keratitis, including fungal and Acanthamoeba keratitis, would be invaluable.

## Data Availability

All data are provided in the paper.

## AUTHOR CONTRIBUTION STATEMENT

Study conceptualisation and design: DSJT, DGS, HSD

Data collection: DSJT, JC, BPG, CSH, LK, AE, ML

Data analysis: DSJT

Data interpretation: DSJT, DGS, HSD

Manuscript drafting: DSJT

Critical revision of manuscript: JC, BPG, CSH, LK, AE, ML, DGS, HSD

Final approval of manuscript: All authors

## Notes

**Funding / Support:** D.S.J.T. acknowledges support from the Medical Research Council / Fight for Sight Clinical Research Fellowship (MR/T001674/1) and the Fight for Sight / John Lee, Royal College of Ophthalmologists Primer Fellowship (24CO4).

**Conflict of interest:** None

### Competing Interest Statement

The authors have declared no competing interest.

### Funding Statement

D.S.J.T. acknowledges support from the Medical Research Council / Fight for Sight Clinical Research Fellowship (MR/T001674/1) and the Fight for Sight / John Lee, Royal College of Ophthalmologists Primer Fellowship (24CO4).

### Author Declarations

The study was approved by the Clinical Governance team in the Nottingham University Hospitals NHS Trust as a clinical audit (Ref: 19-265C).

## REFERENCES

1. Ting DSJ, Ho CS, Deshmukh R, Said DG, Dua HS. Infectious keratitis: an update on epidemiology, causative microorganisms, risk factors, and antimicrobial resistance. Eye (Lond). 2021;35(4):1084–101.

2. Ung L, Bispo PJM, Shanbhag SS, Gilmore MS, Chodosh J. The persistent dilemma of microbial keratitis: Global burden, diagnosis, and antimicrobial resistance. Surv Ophthalmol. 2019;64(3):255–71.

3. Green M, Carnt N, Apel A, Stapleton F. Queensland Microbial Keratitis Database: 2005-2015. Br J Ophthalmol. 2019;103(10):1481–6.

4. Ting DSJ, Ho CS, Cairns J, Elsahn A, Al-Aqaba M, Boswell T, et al. 12-year analysis of incidence, microbiological profiles and in vitro antimicrobial susceptibility of infectious keratitis: the Nottingham Infectious Keratitis Study. Br J Ophthalmol. 2021;105(3):328–33.

5. Khor WB, Prajna VN, Garg P, Mehta JS, Xie L, Liu Z, et al. The Asia Cornea Society Infectious Keratitis Study: A Prospective Multicenter Study of Infectious Keratitis in Asia. Am J Ophthalmol. 2018;195:161–70.

6. Ting DSJ, Ho CS, Cairns J, Gopal BP, Elsahn A, Al-Aqaba M, et al. Seasonal patterns of incidence, demographic factors and microbiological profiles of infectious keratitis: the Nottingham Infectious Keratitis Study. Eye (Lond). 2020; doi: 10.1038/s41433-020-01272-5.

7. Lin L, Duan F, Yang Y, Lou B, Liang L, Lin X. Nine-year analysis of isolated pathogens and antibiotic susceptibilities of microbial keratitis from a large referral eye center in southern China. Infect Drug Resist. 2019;12:1295–302.

8. Shah A, Sachdev A, Coggon D, Hossain P. Geographic variations in microbial keratitis: an analysis of the peer-reviewed literature. Br J Ophthalmol. 2011;95(6):762–7.

9. Tan SZ, Walkden A, Au L, Fullwood C, Hamilton A, Qamruddin A, et al. Twelve-year analysis of microbial keratitis trends at a UK tertiary hospital. Eye (Lond). 2017;31(8):1229–36.

10. Ting DSJ, Settle C, Morgan SJ, Baylis O, Ghosh S. A 10-year analysis of microbiological profiles of microbial keratitis: the North East England Study. Eye (Lond). 2018;32(8):1416–7.

11. Tavassoli S, Nayar G, Darcy K, Grzeda M, Luck J, Williams OM, et al. An 11-year analysis of microbial keratitis in the South West of England using brain-heart infusion broth. Eye (Lond). 2019;33(10):1619–25.

12. Tam ALC, Côté E, Saldanha M, Lichtinger A, Slomovic AR. Bacterial Keratitis in Toronto: A 16-Year Review of the Microorganisms Isolated and the Resistance Patterns Observed. Cornea. 2017;36(12):1528–34.

13. Kowalski RP, Nayyar SV, Romanowski EG, Shanks RMQ, Mammen A, Dhaliwal DK, et al. The Prevalence of Bacteria, Fungi, Viruses, and Acanthamoeba From 3,004 Cases of Keratitis, Endophthalmitis, and Conjunctivitis. Eye Contact Lens. 2020;46(5):265–8.

14. Kaye S, Tuft S, Neal T, Tole D, Leeming J, Figueiredo F, et al. Bacterial susceptibility to topical antimicrobials and clinical outcome in bacterial keratitis. Invest Ophthalmol Vis Sci. 2010;51(1):362–8.

15. Otri AM, Fares U, Al-Aqaba MA, Miri A, Faraj LA, Said DG, et al. Profile of sight-threatening infectious keratitis: a prospective study. Acta Ophthalmologica. 2013;91(7):643–51.

16. Lange C, Feltgen N, Junker B, Schulze-Bonsel K, Bach M. Resolving the clinical acuity categories “hand motion” and “counting fingers” using the Freiburg Visual Acuity Test (FrACT). Graefes Arch Clin Exp Ophthalmol. 2009;247(1):137–42.

17. Grinton M, Sandhu J, Shwe-Tin A, Steel DHW, Ting DSJ. Incidence, characteristics, outcomes and confidence in managing posterior capsular rupture during cataract surgery in the UK: an ophthalmology trainees’ perspective. Eye (Lond). 2021;35(4):1213–20.

18. Chidambaram JD, Venkatesh Prajna N, Srikanthi P, Lanjewar S, Shah M, Elakkiya S, et al. Epidemiology, risk factors, and clinical outcomes in severe microbial keratitis in South India. Ophthalmic Epidemiol. 2018;25(4):297–305.

19. Khoo P, Cabrera-Aguas MP, Nguyen V, Lahra MM, Watson SL. Microbial keratitis in Sydney, Australia: risk factors, patient outcomes, and seasonal variation. Graefes Arch Clin Exp Ophthalmol. 2020;258(8):1745–55.

20. Bourcier T, Thomas F, Borderie V, Chaumeil C, Laroche L. Bacterial keratitis: predisposing factors, clinical and microbiological review of 300 cases. Br J Ophthalmol. 2003;87(7):834–8.

21. Li S, Yi G, Peng H, Li Z, Chen S, Zhong H, et al. How Ocular Surface Microbiota Debuts in Type 2 Diabetes Mellitus. Front Cell Infect Microbiol. 2019;9:202.

22. Jeng BH, Gritz DC, Kumar AB, Holsclaw DS, Porco TC, Smith SD, et al. Epidemiology of ulcerative keratitis in Northern California. Arch Ophthalmol. 2010;128(8):1022–8.

23. Khoo P, Cabrera-Aguas M, Robaei D, Lahra MM, Watson S. Microbial Keratitis and Ocular Surface Disease: A 5-Year Study of the Microbiology, Risk Factors and Clinical Outcomes in Sydney, Australia. Curr Eye Res. 2019;44(11):1195–202.

24. Peng MY, Cevallos V, McLeod SD, Lietman TM, Rose-Nussbaumer J. Bacterial Keratitis: Isolated Organisms and Antibiotic Resistance Patterns in San Francisco. Cornea. 2018;37(1):84–7.

25. Kaliamurthy J, Kalavathy CM, Parmar P, Nelson Jesudasan CA, Thomas PA. Spectrum of bacterial keratitis at a tertiary eye care centre in India. Biomed Res Int. 2013;2013:181564.

26. Ung L, Wang Y, Vangel M, Davies EC, Gardiner M, Bispo PJM, et al. Validation of a Comprehensive Clinical Algorithm for the Assessment and Treatment of Microbial Keratitis. Am J Ophthalmol. 2020;214:97–109.

27. Vital MC, Belloso M, Prager TC, Lanier JD. Classifying the severity of corneal ulcers by using the “1, 2, 3” rule. Cornea. 2007;26(1):16–20.

28. Cariello AJ, Passos RM, Yu MC, Hofling-Lima AL. Microbial keratitis at a referral center in Brazil. Int Ophthalmol. 2011;31(3):197–204.

29. Gopinathan U, Sharma S, Garg P, Rao GN. Review of epidemiological features, microbiological diagnosis and treatment outcome of microbial keratitis: experience of over a decade. Indian J Ophthalmol. 2009;57(4):273–9.

30. Ting DSJ, Bignardi G, Koerner R, Irion LD, Johnson E, Morgan SJ, et al. Polymicrobial Keratitis With Cryptococcus curvatus, Candida parapsilosis, and Stenotrophomonas maltophilia After Penetrating Keratoplasty: A Rare Case Report With Literature Review. Eye Contact Lens. 2019;45(2):e5–e10.

31. Parmar P, Salman A, Kalavathy CM, Kaliamurthy J, Thomas PA, Jesudasan CA. Microbial keratitis at extremes of age. Cornea. 2006;25(2):153–8.

32. Dua HS, Gomes JAP, King AJ, Maharajan VS. The amniotic membrane in ophthalmology. Surv Ophthalmol. 2004;49(1):51–77.

33. Dua HS, Said DG, Messmer EM, Rolando M, Benitez-Del-Castillo JM, Hossain PN, et al. Neurotrophic keratopathy. Prog Retin Eye Res. 2018;66:107–31.

34. Ting DSJ, Rana-Rahman R, Ng JY, Wilkinson DJP, Ah-Kine D, Patel T. Clinical Spectrum and Outcomes of Ocular and Periocular Complications following External-Beam Radiotherapy for Inoperable Malignant Maxillary Sinus Tumors. Ocul Oncol Pathol. 2021;7(1):36–43.

35. Stapleton F, Alves M, Bunya VY, Jalbert I, Lekhanont K, Malet F, et al. TFOS DEWS II Epidemiology Report. Ocul Surf. 2017;15(3):334–65.

36. Ting DSJ, Ghosh S. Acute corneal perforation 1 week following uncomplicated cataract surgery: the implication of undiagnosed dry eye disease and topical NSAIDs. Ther Adv Ophthalmol. 2019;11:2515841419869508.

37. Notara M, Shortt AJ, O’Callaghan AR, Daniels JT. The impact of age on the physical and cellular properties of the human limbal stem cell niche. Age (Dordr). 2013;35(2):289–300.

38. Singhal N, Kumar M, Kanaujia PK, Virdi JS. MALDI-TOF mass spectrometry: an emerging technology for microbial identification and diagnosis. Front Microbiol. 2015;6:791.

39. Ting DSJ, McKenna M, Sadiq SN, Martin J, Mudhar HS, Meeney A, et al. Arthrographis kalrae Keratitis Complicated by Endophthalmitis: A Case Report With Literature Review. Eye Contact Lens. 2020;46(6):e59–e65.

40. Chidambaram JD, Prajna NV, Palepu S, Lanjewar S, Shah M, Elakkiya S, et al. In Vivo Confocal Microscopy Cellular Features of Host and Organism in Bacterial, Fungal, and Acanthamoeba Keratitis. Am J Ophthalmol. 2018;190:24–33.

41. Somerville TF, Corless CE, Sueke H, Neal T, Kaye SB. 16S Ribosomal RNA PCR Versus Conventional Diagnostic Culture in the Investigation of Suspected Bacterial Keratitis. Transl Vis Sci Technol. 2020;9(13):2.

42. Ung L, Bispo PJM, Doan T, Van Gelder RN, Gilmore MS, Lietman T, et al. Clinical metagenomics for infectious corneal ulcers: Rags to riches? Ocul Surf. 2020;18(1):1–12.

43. Ting DSJ, Foo VH, Yang LWY, Sia JT, Ang M, Lin H, et al. Artificial intelligence for anterior segment diseases: Emerging applications in ophthalmology. Br J Ophthalmol. 2021;105(2):158–68.

44. Ting DSJ, Henein C, Said DG, Dua HS. Photoactivated chromophore for infectious keratitis-corneal cross-linking (PACK-CXL): A systematic review and meta-analysis. Ocul Surf. 2019;17(4):624–34.

45. Marasini S, Mugisho OO, Swift S, Read H, Rupenthal ID, Dean SJ, et al. Effect of therapeutic UVC on corneal DNA: Safety assessment for potential keratitis treatment. Ocul Surf. 2021;20:130–8.

46. Kennedy SM, Deshpande P, Gallagher AG, Horsburgh MJ, Allison HE, Kaye SB, et al. Antimicrobial Activity of Poly-epsilon-lysine Peptide Hydrogels Against Pseudomonas aeruginosa. Invest Ophthalmol Vis Sci. 2020;61(10):18.

47. Mayandi V, Xi Q, Leng Goh ET, Koh SK, Jie Toh TY, Barathi VA, et al. Rational Substitution of ε-Lysine for α-Lysine Enhances the Cell and Membrane Selectivity of Pore-Forming Melittin. J Med Chem. 2020;63(7):3522–37.

48. Ting DSJ, Beuerman RW, Dua HS, Lakshminarayanan R, Mohammed I. Strategies in Translating the Therapeutic Potentials of Host Defense Peptides. Front Immunol. 2020;11:983.

