## Supplementary material for "Risk Factors, Clinical Outcomes and Prognostic Factors of Bacterial Keratitis: The Nottingham Infectious Keratitis Study": Table 1

**Table 1.** Summary of the demographic factors, risk factors and baseline clinical characteristics of bacterial keratitis presented to Queen’s Medical Centre, Nottingham, UK. Comparison between culture-proven and culture-negative cases are performed.

| **Parameters** | **All cases**  **Total N = 283;**  **N (%)** | **Culture-proven**  **Total N = 128;**  **N (%)** | **Culture-negative**  **Total N = 155;**  **N (%)** | **P-value**^#^ |
| --- | --- | --- | --- | --- |
| Age, years  Gender  Female  Male  Laterality  Left  Right  Risk factors^$^  OSD*  Contact lens wear  Immunosuppression**  Prior corneal surgery  Topical corticosteroids  Trauma  None identified  Presenting CDVA, in logMAR  0.0 – 0.3  <0.3 – 0.6  <0.6-1.0  <1.0  Size of epithelial defect  Small (≤3mm)  Moderate (3.1-6mm)  Large (>6mm)  Size of infiltrate  Small (≤3mm)  Moderate (3.1-6mm)  Large (>6mm)  Location  Central  Paracentral  Peripheral  Hypopyon  Yes  No  Hospitalisation required  Yes  No  Duration of hospitalisation, days | 54.4 ± 21.0  139 (49.1)  144 (50.9)  145 (51.2)  138 (48.8)  134 (47.3)  100 (35.3)  52 (18.4)  39 (13.8)  31 (11.0)  25 (8.8)  10 (3.5)  89 (31.4)  41 (14.5)  29 (10.2)  124 (43.8)  172 (60.8)  63 (22.3)  48 (17.0)  183 (64.7)  62 (21.9)  38 (13.4)  110 (38.9)  106 (37.5)  67 (23.7)  82 (29.0)  201 (71.0)  162 (57.2)  121 (42.8)  8.0 ± 8.3 | 58.5 ± 21.3  61 (47.7)  67 (52.3)  72 (56.3)  56 (43.7)  59 (46.1)  41 (32.0)  27 (21.1)  25 (19.5)  21 (16.4)  10 (7.8)  3 (2.3)  20 (15.6)  20 (15.6)  15 (11.7)  73 (57.0)  58 (45.3)  43 (33.6)  27 (21.1)  65 (50.8)  40 (31.2)  23 (18.0)  57 (44.5)  53 (41.4)  18 (14.1)  60 (46.9)  68 (53.1)  95 (74.2)  33 (25.8)  8.8 ± 9.2 | 51.1 ± 20.1  78 (50.3)  77 (49.7)  73 (47.1)  82 (52.9)  75 (48.4)  59 (38.1)  25 (16.1)  14 (9.0)  10 (6.5)  15 (9.7)  7 (4.5)  69 (44.5)  21 (13.5)  14 (9.0)  51 (32.9)  114 (73.5)  20 (12.9)  21 (13.5)  118 (76.1)  22 (14.2)  15 (9.7)  53 (34.2)  53 (34.2)  49 (31.6)  22 (14.2)  133 (85.8)  67 (43.2)  88 (56.8)  6.0 ± 4.9 | 0.004  0.66  0.13  0.030  0.70  0.29  0.28  0.011  0.008  0.58  0.32  <0.001  <0.001  <0.001  0.002  <0.001  <0.001  0.06 |

OSD = Ocular surface disease; CDVA = Corrected-distance-visual-acuity

Continuous values are presented as mean ± standard deviation (SD).

^$^Some patients had more than 1 risk factor identified.

*Includes dry eye disease, meibomian gland disease, neurotrophic keratopathy, exposure keratopathy, previous corneal infection, corneal erosion syndrome, limbal stem cell deficiency, cicatricial conjunctivitis, band keratopathy, and bullous keratopathy.

**Includes diabetes, use of systemic immunosuppressive drugs, malnutrition, and immunodeficiency.

^#^Comparison between culture-positive and culture-negative cases. Chi-square and unpaired T-test were used for categorical and continuous variables, respectively. Significant values are underlined.
