## Supplementary material for "Risk Factors, Clinical Outcomes and Prognostic Factors of Bacterial Keratitis: The Nottingham Infectious Keratitis Study": Table 2

**Table 2.** Summary of risk factors based on different age groups.

| **Risk factors** | **Age ≤ 50 years**  **Total N = 118**  **N (%)** | **Age > 50 years**  **Total N = 165**  **N (%)** | **P-value** |
| --- | --- | --- | --- |
| Presence of risk factors  None  One  Two  Three or more  Type of risk factors  OSD*  Contact lens wear  Immunosuppression**  Prior corneal surgery  Topical corticosteroids  Trauma | 4 (3.4)  78 (66.1)  26 (22.0)  10 (8.5)  54 (45.8)  68 (57.6)  9 (7.6)  15 (12.7)  8 (6.8)  10 (8.5) | 6 (3.6)  111 (67.3)  40 (24.2)  8 (4.8)  80 (50.6)  32 (19.4)  43 (26.1)  24 (14.5)  23 (14.6)  15 (9.1) | 0.66  <0.001  0.65  <0.001  <0.001  0.19  0.06  0.86 |

OSD = Ocular surface disease

*Includes dry eye disease, meibomian gland disease, neurotrophic keratopathy, exposure keratopathy, previous corneal infection, corneal erosion syndrome, limbal stem cell deficiency, cicatricial conjunctivitis, band keratopathy, and bullous keratopathy.

**Includes diabetes, use of systemic immunosuppressive drugs, malnutrition, and immunodeficiency
