## Supplementary material for "Risk Factors, Clinical Outcomes and Prognostic Factors of Bacterial Keratitis: The Nottingham Infectious Keratitis Study": Table 3

**Table 3.** Summary of causative organisms of bacterial keratitis in Nottingham, UK, and their association with different risk factors.

|  | Total*  Total N=138  N (%) | OSD  Total N=63  N (%) | CL wear  Total N=45  N (%) | CS  Total N=25  N (%) | TC  Total N=21  N (%) | P-value** |
| --- | --- | --- | --- | --- | --- | --- |
| Gram-positive  Staphylococci  Streptococci  Other GP  Gram-negative | 70 (50.7)  36 (26.1)  16 (11.6)  18 (13.0)  68 (49.3) | 38 (60.3)  20 (31.7)  9 (14.3)  9 (14.3)  25 (39.7) | 15 (33.3)  9 (20.0)  2 (4.4)  4 (8.9)  30 (66.7) | 16 (64.0)  10 (40.0)  4 (16.0)  2 (8.0)  9 (36.0) | 13 (61.9)  7 (33.3)  5 (23.8)  2 (9.5)  8 (38.1) | 0.017 |
| Pseudomonas  Moraxella  Other GN | 44 (31.9)  14 (10.1)  10 (7.2) | 13 (20.6)  8 (12.7)  4 (6.3) | 23 (51.1)  3 (6.7)  4 (8.9) | 6 (24.0)  3 (12.0)  0 (0.0) | 4 (19.0)  3 (14.3)  1 (4.8) |  |

OSD = Ocular surface disease; CL = Contact lens; TC = Topical corticosteroids; CS = Previous corneal surgery; GP = Gram-positive; GN = Gram-negative

*The total number of organisms exceeded the total number of culture-positive cases as some cases were polymicrobial. In addition, some cases had more than one risk factor identified, and the same implicated organism was included in more than one group of risk factor.

**Comparison of the causative organisms among different risk factors were performed. The analysis was performed at the level of Gram-positive versus Gram-negative bacteria only.
