## Supplementary material for "Risk Factors, Clinical Outcomes and Prognostic Factors of Bacterial Keratitis: The Nottingham Infectious Keratitis Study": Table 4

**Table 4.** Prognostic factors for poor visual outcome [defined as corrected-distance-visual-acuity (CDVA) of <0.60 logMAR] and poor corneal healing (defined as >30 days to achieve complete healing or occurrence of corneal perforation or uncontrolled infection) in bacterial keratitis.

|  | **Poor visual outcome** | | **Poor corneal healing** | |
| --- | --- | --- | --- | --- |
| **Parameters** | **Odd ratio (95% CI)** | **P-value*** | **Odd ratio (95% CI)** | **P-value*** |
| Age > 50 years  Female gender  Right eye  Epithelial defect size >3mm  Infiltrate size >3mm  Central ulcer  Presence of hypopyon  Positive culture results  Presenting CDVA <0.6 | 2.61 (1.24 – 5.47)  0.81 (0.41 – 1.62)  0.81 (0.41 – 1.60)  0.69 (0.19 – 2.50)  4.07 (1.21 – 13.73)  2.13 (1.01 – 4.51)  0.47 (0.21 – 1.08)  1.17 (0.56 – 2.46)  29.70 (10.47-84.18) | 0.011  0.56  0.54  0.57  0.024  0.047  0.08  0.67  <0.001 | 1.86 (1.06 – 3.24)  1.27 (0.73 – 2.19)  1.82 (1.05 – 3.16)  1.23 (0.44 – 3.44)  3.46 (1.24 – 9.70)  1.30 (0.67 – 2.54)  1.02 (0.50 – 2.07)  1.15 (0.63 – 2.10)  2.22 (1.19 – 4.15) | 0.030  0.40  0.033  0.69  0.018  0.43  0.96  0.65  0.013 |

*Multivariable logistic regression analysis was performed. Significant p-values are underlined.
